# The Characteristics of Health Care Workers with COVID-19 and Relationship Between COVID-19 Mortality and BCG/Tuberculosis History: a multi-center study

**DOI:** 10.1101/2020.10.08.20209403

**Authors:** Ş Torun, Ş Özkaya, N Şen, F Kanat, I Karaman, Ş Yosunkaya, Ö Ş Dikiş, A Asan, S A Eroğlu, S S Atal, Ö Ayten, N Aksel, H Ermiş, N Özçelik, M Demirelli, İ Kara, Ş Sümer, K Marakoğlu, F Üzer, Y Uyar, T Çiçek, Z E Ünsal, H Vatansev, B B Yıldırım, T Kuruoğlu, A Atilla, Y Ersoy, B Kandemir, Y Durduran, F G Cihan, N Demirbaş, F Yıldırım, M S Akcay

## Abstract

**Background:** Today, COVID-19 pandemic has brought countries’ health services into sharp focus. Despite the low incidence of cases(1.2%) and high mortality rate(2.4%) among Turkish population, the low mortality rate(0.3%) despite the high incidence(11.5%) declared in healthcare workers drew our group’s attention. Therefore, we aimed to report the characteristics of infected health-care workers and investigate the relationship between BCG vaccine and tuberculosis history with COVID-19 mortality in infected health-care worker population.

**Method:** This study was conducted in three hospitals to assess the clinical presentations, disease severity and correlation with BCG vaccine and tuberculous history in COVID-19 positive health-care workers by an online questionnaire platform. The relationship between characteristics and tuberculosis history were investigated according to hospitalization status of the patients.

**Result:** Total of 465 infected healthcare workers included in the study. The rate of history of direct care and contact to tuberculosis patient, presence of previous tuberculosis treatment and BCG scar, presence of radiological infiltrations was significantly higher in hospitalized healthcare workers. The ratio of direct care and direct contact to the patient with tuberculosis, and presence of family history of tuberculosis were statistically significantly higher in patients with radiological infiltrations.

**Conclusion:** Although COVID-19 risk and incidence are higher among healthcare workers compared to the normal population due to higher virus load, this study brings evidence for the fact that the lower mortality rate seen in infected healthcare workers might be due to healthcare workers’ frequent exposure to tuberculosis bacillus and the mortality-reducing effects of BCG vaccine, despite the higher hospitalization rate and radiological infiltrations due to over-triggered immune system.

## Introduction

The novel coronavirus disease 2019 (COVID-19) was first reported in Wuhan, China, in late December 2019. (1,2). On 30 January 2020, the World Health Organization (WHO) declared a public health emergency for novel COVID□19 disease, which is caused by severe acute respiratory syndrome coronavirus 2 (SARS□CoV□2). Since then, the entire world is battling with COVID□19 pneumonia that can be dreadfully lethal in high□risk population. One of the most concerning aspects of COVID-19 is the risk for infection among frontline healthcare workers.

Since the beginning of the pandemic, several statistical analysis and reports were published to prove the hypothesis about potential protective role of BCG vaccine against COVID-19. Data regarding number of COVID-19 patients from countries with universal BCG vaccination and those who discontinued vaccination showed that the disease mortality is reduced in countries with universal BCG (3,4). As of September 12 2020, the mortality rate in Turkey was reported as 2.4%, compared to 3.1% global mortality rate (2). According to the recent statement by the Ministry of Health of Turkish Republic, the current mortality rate for healthcare workers has been declared as 0.3%. Despite the low incidence of cases(1.2%) and high mortality rate(2.4%) among public, the low mortality rate(0.3%) despite the high incidence(11.5%) declared in healthcare workers in Turkey drew our group’s attention. Also, data regarding the characteristics of front-line health-care workers and their risk of COVID-19 is limited. Therefore, we aimed to report the characteristics of infected health-care workers and investigate the relationship between BCG vaccine and tuberculosis history with COVID-19 mortality in infected health-care worker population.

## Material and Methods

The online platform of Google documents was used as a platform to create online questionnaires that were automatically hosted via a unique URL. Password protected access to the URL link and a unique study ID gave patients around the clock access from anywhere in the Turkey. Unique study ID ensured confidentially of all self-reported data. Patient responses were secured using a “Cloud” database where the data was automatically sorted, scaled and scored by custom Excel formulas. Researchers downloaded real-time questionnaire responses in multiple formats (e.g. excel) which was then analyzed with a statistical software of choice. This study was conducted in three hospitals to assess the clinical presentations, disease severity and correlation with BCG vaccine and tuberculous history in COVID-19 positive health-care workers. Confidentiality of the study participants’ information was maintained throughout the study by making the participants’ information anonymous and asking the participants to provide honest answers. Informed consent was obtained from each participant prior to participation. This protocol approved by the applicable IRBs/ECs [institutional review boards/ethical committees of Baskent University, Ankara, Turkey; approval number: KA20/171-20/64, July 1, 2020] with respect to its scientific content.

## Results

Total of 465 infected healthcare workers included in the study. Demographic characteristics of the patients according to their hospitalization status was shown in Table I. 58.9% of the patients who were hospitalized and 53.9% of the patients who were not hospitalized were women, and the differences between two groups were not statistically significant (p> 0.05). The mean age of the non-hospitalized patients (34.66 ± 8.93) was lower than the mean age of the hospitalized patients (39.59 ± 46.17), however the difference was not statistically significant (p> 0.05). Working time in the unit was statistically significantly higher in the hospitalized group (p <0.05). Chronic disease was present in 18.1% of patients who were not hospitalized and 28.7% of patients who were hospitalized, and the differences between groups were statistically significant (p <0.05).

**Table I.** Demographic characteristics of the patients according to their hospitalization status.

|  | Covid-19 Hospitalization |  | p |
| --- | --- | --- | --- |
|  | No<br>(n=248; %53.3) | Yes<br>(n=217; %46.7) |  |
| **Gender, n (%)** |  |  |  |
| Female | 146 (58.9) | 117 (53.9) | 0.282<sup>a</sup> |
| Male | 102 (41.1) | 100 (46.1) |  |
| **Age**(Mean ± SD) | 34.66±8.93 | 39.59±46.17 | 0.056<sup>b</sup> |
| **Working time in the unit**(Mean ± SD) | 6.42±5.64 | 8.15±7.49 | 0.046<sup>b</sup> |
| **Working place, n (%)** |  |  |  |
| Inpatient service | 63 (25.4) | 76 (35.0) | 0.097<sup>a</sup> |
| Outpatient clinic | 29 (11.7) | 27 (12.4) |  |
| Intensive care unit | 33 (13.3) | 34 (15.7) |  |
| Emergency | 16 (6.5) | 13 (6.0) |  |
| Other | 34 (13.7) | 25 (11.5) |  |
| More than one unit | 73 (29.4) | 42 (19.4) |  |
| **Job, n (%)** |  |  |  |
| Doctor | 92 (37.1) | 70 (32.3) | 0.441<sup>a</sup> |
| Nurse | 95 (38.3) | 84 (38.7) |  |
| Other | 61 (24.6) | 63 (29.0) |  |
| **Presence of chronic diseases, n (%)** |  |  |  |
| No | 203 (81.9) | 154 (71.3) | 0.007<sup>a</sup> |
| Yes | 45 (18.1) | 62 (28.7) |  |

a. Chi-square test, b. Mann Whitney U Test, SD: Standard Deviation

The relationship between tuberculosis history and characteristics of the patients according to hospitalization status was described in Table II. 64.5% of the patients who did not receive hospitalization and 66.0% of the patients who were hospitalized had a history of direct care to a tuberculosis patient, and the difference according to hospitalization status was not statistically significant (p> 0.05). A tuberculosis unit in the institution was present in 58.4% of patients who did not receive hospitalization, and 53.0% of patients who were hospitalized, and the difference was not significant (p> 0.05). The rate of direct care for patients with tuberculosis was 24.7% in non-hospitalized patients and 35.5% in hospitalized patients, and the differences between groups were statistically significant (p <0.05). Direct contact with tuberculosis patients was found in 27.6% of non-hospitalized patients and 37.2% of hospitalized patients, and the differences between groups were statistically significant (p <0.05). 6.5% of the hospitalized patients had previously received tuberculosis treatment (p <0.05). BCG scar was statistically significantly higher in patients with hospitalization, since 216 of the patients with scar were not hospitalized and 205 were hospitalized (p <0.05). The presence of radiological pneumonia was statistically significantly higher in hospitalized patients (p <0.05).

**Table II.**
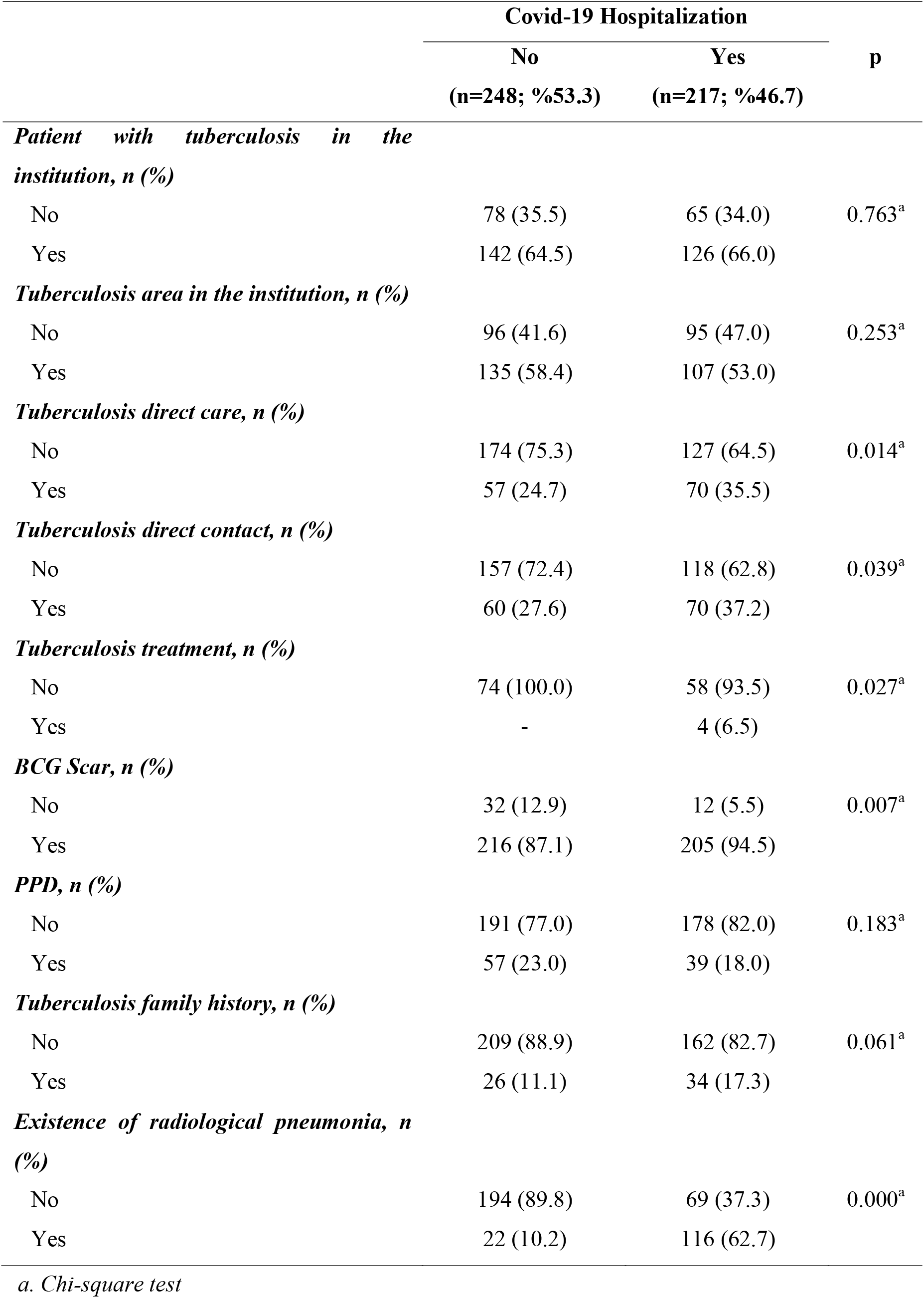
The relationship between tuberculosis history and characteristics of the patients according to hospitalization status

|  | Covid-19 Hospitalization |  | p |
| --- | --- | --- | --- |
|  | No<br>(n=248; %53.3) | Yes<br>(n=217; %46.7) |  |
| *Patient with tuberculosis in the institution, n (%)* |  |  |  |
| No | 78 (35.5) | 65 (34.0) | 0.763<sup>a</sup> |
| Yes | 142 (64.5) | 126 (66.0) |  |
| *Tuberculosis area in the institution, n (%)* |  |  |  |
| No | 96 (41.6) | 95 (47.0) | 0.253<sup>a</sup> |
| Yes | 135 (58.4) | 107 (53.0) |  |
| *Tuberculosis direct care, n (%)* |  |  |  |
| No | 174 (75.3) | 127 (64.5) | 0.014<sup>a</sup> |
| Yes | 57 (24.7) | 70 (35.5) |  |
| *Tuberculosis direct contact, n (%)* |  |  |  |
| No | 157 (72.4) | 118 (62.8) | 0.039<sup>a</sup> |
| Yes | 60 (27.6) | 70 (37.2) |  |
| *Tuberculosis treatment, n (%)* |  |  |  |
| No | 74 (100.0) | 58 (93.5) | 0.027<sup>a</sup> |
| Yes | - | 4 (6.5) |  |
| *BCG Scar, n (%)* |  |  |  |
| No | 32 (12.9) | 12 (5.5) | 0.007<sup>a</sup> |
| Yes | 216 (87.1) | 205 (94.5) |  |
| *PPD, n (%)* |  |  |  |
| No | 191 (77.0) | 178 (82.0) | 0.183<sup>a</sup> |
| Yes | 57 (23.0) | 39 (18.0) |  |
| *Tuberculosis family history, n (%)* |  |  |  |
| No | 209 (88.9) | 162 (82.7) | 0.061<sup>a</sup> |
| Yes | 26 (11.1) | 34 (17.3) |  |
| *Existence of radiological pneumonia, n (%)* |  |  |  |
| No | 194 (89.8) | 69 (37.3) | 0.000<sup>a</sup> |
| Yes | 22 (10.2) | 116 (62.7) |  |

a. Chi-square test

According to the radiological pneumonia status (Table III), the differences between the hospitalized and non-hospitalized groups in terms of care to patients with tuberculosis in the institution, presence of tuberculosis unit in the institution, tuberculosis treatment history status, BCG scar, PPD, BCG scar number and PPD results were not statistically significant (p> 0.05). On the other hand, the ratio of direct care to the patient with tuberculosis, direct contact with the patient with tuberculosis, and presence of family history of tuberculosis were statistically significantly higher in the group with radiological pneumonia (p <0.05). We noted that the only one(0.2%) healthcare worker died from COVID-19 pneumonia.

**Table III.** The relationship between tuberculosis history and characteristics of the patients according to existence of radiological pneumonia in patients

|  | Radiological pneumonia* |  |  |
| --- | --- | --- | --- |
|  | No | Yes | p |
|  | (n=263; %65.6) | (n=138; %29.7) |  |
| *Patient with tuberculosis in the institution, n (%)* |  |  |  |
| No | 81 (35.1) | 39 (32.5) | 0.631<sup>a</sup> |
| Yes | 150 (64.9) | 81 (67.5) |  |
| *Tuberculosis area in the institution, n (%)* |  |  |  |
| No | 113 (45.9) | 62 (47.0) | 0.847<sup>a</sup> |
| Yes | 133 (54.1) | 70 (53.0) |  |
| *Tuberculosis direct care, n (%)* |  |  |  |
| No | 175 (72.9) | 79 (60.8) | 0.016<sup>a</sup> |
| Yes | 65 (27.1) | 51 (39.2) |  |
| *Tuberculosis direct contact, n (%)* |  |  |  |
| No | 153 (69.5) | 73 (57.9) | 0.029<sup>a</sup> |
| Yes | 67 (30.5) | 53 (42.1) |  |
| *Tuberculosis treatment, n (%)* |  |  |  |
| No | 71 (98.6) | 40 (93.0) | 0.114<sup>a</sup> |
| Yes | 1 (1.4) | 3 (7.0) |  |
| *BCG Scar, n (%)* |  |  |  |
| No | 31 (11.8) | 8 (5.8) | 0.054<sup>a</sup> |
| Yes | 232 (88.2) | 130 (94.2) |  |
| *PPD, n (%)* |  |  |  |
| No | 205 (77.9) | 108 (78.3) | 0.942<sup>a</sup> |
| Yes | 58 (22.1) | 30 (21.7) |  |
| *Tuberculosis family history, n (%)* |  |  |  |
| No | 221 (89.5) | 97 (77.6) | 0.002<sup>a</sup> |
| Yes | 26 (10.5) | 28 (22.4) |  |
| Number of BCG Scar, Average ± SD | 1.37±0.64 | 1.46±0.65 | 0.079<sup>b</sup> |
| PPD Result, Average ± SD | 9.51±7.87 | 10.31±9.33 | 0.835<sup>b</sup> |

*\*Since radiological pneumonia information was not available in 13.8% of the patients, the rates are given according to the ones whose information can be accessed.*

## Discussion

Today, COVID-19 pandemic has brought countries’ health services into sharp focus. While governments move to slow the spread of the virus, health workers, especially women, are on the front lines of the epidemic. Health-care workers are at increased risk of being exposed to SARS-CoV-2 and could potentially have a role in hospital transmission. Turkey is one of the largest populations with health workers as employing 173 health care workers per 10,000 population. After the first case announced on March 11, until September 12, total of 29,065,028 patients and 291,162 patients were documented in all over the world and in Turkey, respectively. The global mortality rate was reported as 3.1%(926,283 patients) compared to 2.4%(7056 patients). in Turkey.

According to the recent statement by the Ministry of Health of Turkish Republic, approximately 30.000 health workers were infected with SARS-CoV-2 and approximately 100 employees were died of COVID-19, including 33 doctors and 66 other health workers in Turkey. Therefore, the current mortality rate in Turkey for healthcare workers has been declared as 0.3%. In our study, among 465 infected healthcare workers, only 1(0.2%) healthcare worker died from COVID-19. Despite the high number of cases and mortality rate among public, the low mortality rate declared in healthcare workers drew our group’s attention. We thought that the low mortality rate can be explained by the fact that healthcare workers are exposed more frequently to bacterial and viral agents such as *Mycobacterium tuberculosis* than the society, therefore, their immune system are constantly active. Supportively, in our study, contact with tuberculosis and the history of BCG were investigated and exposure was noted over 80s%.

A study with 2.135.190 people in the UK and USA using the COVID-19 Symptom Study app between March-April, 2020 noted that front-line health-care workers had at least a three-fold increased risk of reporting a positive COVID-19 test and COVID-19 infection, compared with the general community. Also, public health authorities suggested that 10–20% of SARS-CoV-2 infections occur among health-care workers (5,6). However, the positivity in screening results of asymptomatic healthcare workers has been reported lower than this rate. The rate of asymptomatic infections among healthcare workers was reported as 3% in the United Kingdom (UK) (7). In a study from the Netherlands, after screening with PCR, 1% of health care workers were found as infected with SARS-CoV-2 (8). An early, single□center, retrospective analysis from Zhongnan Hospital, Wuhan, of 138 patients with COVID□19 pneumonia showed that 41% of infections were thought to be hospital□acquired. Approximately 70% of these patients were healthcare workers, with one individual requiring intensive care and zero deaths. The study showed that infected hospital staff worked mostly on general wards (77.5%) followed by the emergency department (17.5%) and critical care (5%) (9). A study of more than 72,000 patients with COVID-19 by the Chinese Centre for Disease Control and Prevention showed that by early February around 3000 healthcare workers had become infected, accounting for 3.8% of all cases of COVID-19. There were five deaths in this cohort giving a case fatality ratio of 0.3%; this is approximately one□sixth of the overall case fatality rate(2.3%) in the study, half of which occurred in patients aged 20–59 years (0.6%). In that study, early January and through February 2020, the proportion of confirmed cases in healthcare workers graded severe or critical decreased from 45% to 8.7% (10). According to Ministry of Health of Turkish Republic, 29865 health workers were infected, while the rate of getting the infection for the whole society was 1.2%, this rate reached 11.5% for healthcare workers.

In this study, the rate of history of direct care and direct contact to tuberculosis patient, previous tuberculosis treatment, presence of BCG scar, was significantly higher in hospitalized healthcare workers. The presence of radiological infiltrations was also significantly higher in hospitalized group. In the group with radiological infiltrations, the ratio of direct care to the patient with tuberculosis, direct contact with the patient with tuberculosis, and presence of family history of tuberculosis were statistically significantly higher. Therefore, we concluded that a history of tuberculosis contact might worsen the clinical picture of COVID-19 course by triggering immune system and causing further radiological infiltrations, however, it also decreases the mortality rate. To the authors’ knowledge, this is the first clinical study that investigates the characteristics of infected healthcare workers in Turkey. However, we are aware that this study is limited to fully support the beforementioned conclusion. Further in-depth studies are needed to provide evidence for such relation.

In conclusion, although COVID-19 risk and incidence are higher among healthcare workers compared to the normal population due to higher virus load, this study brings evidence for the fact that the lower mortality rate seen in infected healthcare workers might be due to healthcare workers’ frequent exposure to tuberculosis bacillus and the mortality-reducing effects of BCG vaccines. Further descriptive research can enable the conduction of larger studies that might document this potential association.

## Data Availability

The research article data that used to support the findings of this study is available from the corresponding author upon request.

## Acknowledgments

None.

## Author Contributions

Guarantors of integrity of entire study; all authors, Study concepts/study design or data acquisition or data analysis/interpretation; all authors; manuscript drafting or manuscript revision for important intellectual content; all authors; approval of final version of submitted manuscript; all authors.

## Declaration of Competing Interest

All authors have contributed significantly, and that all authors are in agreement with the content and honesty of the manuscript. All authors declare that they have no conflict of interest.

## Notes

### Competing Interest Statement

The authors have declared no competing interest.

### Author Declarations

This protocol approved by the applicable IRBs/ECs [institutional review boards/ethical committees of Baskent University, Ankara, Turkey; approval number: KA20/171-20/64, July 1, 2020] with respect to its scientific content.

